# INTERLEUKIN-1 RECEPTOR ANTAGONIST LEVELS IN PATIENTS WITH HEART FAILURE AND REDUCED EJECTION FRACTION TREATED WITH ANAKINRA

**DOI:** 10.64898/2026.04.17.26351024

**Authors:** Jazmin Kelly, Eleonora Mezzaroma, Andrea Roscioni, Chantel McSkimming, Adolfo Mauro, Pratyush Narayan, Michele Golino, Cory Trankle, Justin M. Canada, Stefano Toldo, Benjamin W. Van Tassell, Antonio Abbate

## Abstract

**Background:** Patients with heart failure and reduced ejection fraction (HFrEF) commonly show signs of systemic inflammation. Interleukin-1 (IL-1) is a pro-inflammatory cytokine, known to modulate cardiac function. We aimed to determine the effects of treatment with anakinra, recombinant IL-1 receptor antagonist (IL-1Ra), on plasma IL-1Ra levels.

**Methods:** We measured IL-1Ra levels at baseline and longest available follow-up to 24 weeks in 63 patients (44 males, 40 self-identified Black-Americans) with recent hospitalization for HFrEF, and systemic inflammation (C-reactive protein [CRP] levels >2 mg/L) who were assigned to anakinra (n=42 [66.7%]) or placebo (n=21 [33.3%]) as part of the REDHART2 clinical trial (NCT0014686). Cardiorespiratory fitness was measured as peak oxygen consumption (VO_2peak_).

**Results:** Baseline plasma IL-1Ra levels were 380 [290 to 1046] pg/mL. On-treatment IL-1Ra levels were significantly higher in the patients treated with anakinra vs. placebo (3,994 [3,372 to 5,000] pg/mL vs. 492 [304 to 1370] pg/mL, *P*<0.001). The longest available follow-up was 6 weeks in 10 patients (15.9%), 12 weeks in 12 patients (19%), and 24 weeks in 41 patients (65.1%). On-treatment IL-1Ra levels and interval change in IL-1Ra showed a modest inverse correlation with on-treatment CRP levels (R=-0.269, *P*=0.033 and R=-0.355, *P*=0.004, respectively) and no statistically significant correlations with VO_2peak_ values (*P*>0.05).

**Conclusions:** Patients with recently decompensated HFrEF and systemic inflammation treated with recombinant IL-1Ra, anakinra, have a significant several-fold increase in plasma IL-1Ra levels. On-treatment IL-1Ra levels however show only a modest correlation with CRP levels and not with (VO_2peak_).

## INTRODUCTION

Heart failure with reduced ejection fraction (HFrEF) is increasingly recognized as a syndrome of impaired cardiac function associated with not only neurohormonal activation but also by a systemic inflammatory response contributing to disease progression and functional limitation.^1^ Early experimental studies have demonstrated that Interleukin-1 (IL-1), an apical pro-inflammatory cytokine, exerts direct cardiodepressant effects and impairs β-adrenergic responsiveness, contributing to reduced cardiac performance, exercise intolerance and supporting a mechanistic role for inflammation, and IL-1, in heart failure pathophysiology. ^2-3^

On this basis, pharmacologic inhibition of IL-1 signaling is being investigated as a therapeutic strategy in heart failure. IL-1 blockade with canakinumab in patients with prior myocardial infarction reduced both recurrent atherothrombotic events and heart failure-related events^4^. Patients with significantly lower event rates were those who achieved inflammation resolution, as shown by on-treatment C-reactive protein (CRP) <2 mg/L.^5-6^ In patients with HFrEF, treated with canakinumab or recombinant human IL-1 receptor antagonist (IL-1Ra), anakinra, reduced systemic inflammation, measured as CRP and was associated with an improved cardiorespiratory fitness, measured a peak oxygen consumption (VO_2peak_), ^7-9^ providing proof-of-concept that targeting IL-1 may favorably influence functional capacity in patients with HFrEF.^10^ However, in a recent clinical trial of anakinra or placebo in patients with recently decompensated HFrEF enrolled within 14 days of discharge and followed for 24 weeks, CRP levels improved in both the anakinra and placebo groups, with only a marginal greater reduction in the anakinra arm, and CRP levels remained elevated (≥2 mg/L) in a substantial proportion of treated patients, suggesting incomplete inflammatory suppression.^11^ Anakinra also failed to significantly improve VO_2peak_ compared with placebo.^11^ Nevertheless, patients with inflammation resolution, as shown by CRP <2 mg/L, had favorable changes in VO_2peak_ and outcomes.^11^ These findings may indicate that incomplete inhibition of IL-1 signaling with anakinra may be contributing to the neutral functional outcome observed in REDHART2. Since anakinra competitively blocks the IL-1 receptor, we aimed to determine whether IL-1Ra concentrations in patients treated with anakinra were associated with reductions in systemic inflammation and improvements in cardiorespiratory fitness.

## METHODS

The protocol and the data that support the findings of this study are available from the corresponding author upon reasonable request.

### Study design

This study is a secondary pharmacokinetic and biomarker analysis conducted within the REcently Decompensated Heart failure Anakinra Response Trial 2 (REDHART2; ClinicalTrials.gov identifier: NCT03797001), a single-center, randomized, double-blind, phase II clinical trial with a 2:1 allocation to anakinra or placebo in patients with recently decompensated HFrEF, enrolled within 14 days of hospital discharge and treated for 24 weeks.^11-12^ For this analysis, paired plasma samples suitable for IL-1Ra measurement were available from 63 participants, including 42 patients treated with anakinra and 21 receiving placebo (**Figure 1**).

**Figure 1.**
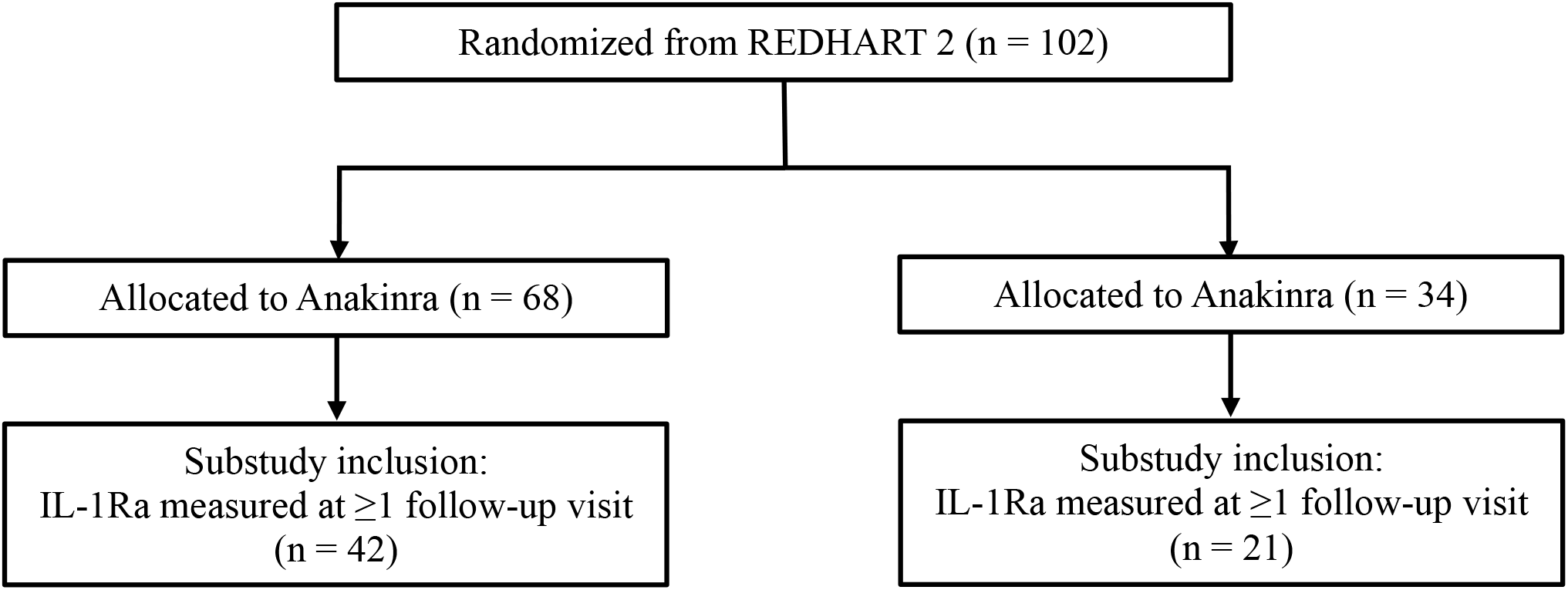
Screening and enrollment of patients. The Consolidated Standards of Reporting Trials (CONSORT) diagram reflects the derivation of the study population from the parent REDHART2 trial. A total of 102 patients were randomized to anakinra or placebo in a 2:1 ratio. Of these, 63 patients had IL-1Ra levels measured during at least one follow-up visit and were included in the present substudy, including 42 in the anakinra group and 21 in the placebo group. The remaining 39 patients were not included due to the absence of IL-1Ra measurements at follow-up.

### Laboratory data

Laboratory methods for biomarker collection and processing in the REDHART2 trial have been described previously.^11-12^ Blood samples were obtained at baseline and during follow-up visits, with measurements performed at the longest available time point during treatment, up to 24 weeks. Plasma samples were analyzed using enzyme-linked immunosorbent assay (Quantikine ELISA Kit, R&D Systems) to measure human interleukin-1Ra (Human IL-1Ra/IL-1F3) following the manufacturer’s instructions. Changes in IL-1Ra concentrations from baseline to follow-up were evaluated in relation to changes in CRP levels to assess biological exposure to IL-1 blockade.

### Clinical data

The clinical characterization of patients in the REDHART2 trial has been described previously.^11-12^ Briefly, clinical data were obtained through structured assessments and medical record review, together with laboratory testing, transthoracic echocardiography, and cardiopulmonary exercise testing performed at baseline and scheduled follow-up visits. Changes in IL-1Ra concentrations from baseline to follow up were evaluated in relation to changes in VO_2peak_ and cardiac function to assess biological exposure to IL-1 blockade.^11-12^

### Statistical analysis

Statistical methods applied in the REDHART2 trial have been described previously.^11-12^ Descriptive statistics were used to summarize baseline and clinical characteristics of the study population. Continuous variables are presented as median [interquartile range]. Comparisons of continuous variables between independent groups were performed using the Mann–Whitney U test, whereas paired comparisons between baseline and follow-up measurements were conducted using the Wilcoxon signed-rank test. Changes from baseline were calculated as follow-up minus baseline values, with positive values indicating an increase and negative values indicating a decrease. Analyses were performed using available observations and missing data were not imputed. All statistical analyses were performed using SPSS software, version 31.0 (IBM SPSS Statistics, Chicago, IL), with statistical significance defined as a two-sided *P*-value <0.05.

### Regulatory data

All methods were carried out in accordance with relevant guidelines and regulations. All experimental protocols were approved by the Virginia Commonwealth University Institutional Review Board. All subjects provided written informed consent to be part of the trial in accordance with the Virginia Commonwealth University Institutional Review Board (protocol number HM20014686).

## RESULTS

### Characteristics of the Patients

The clinical characteristics of the 63 patients are summarized in **Table 1**. Patients were predominantly male (44, 70%), and self-identified as Black Americans (40, 64%), with a median age of 61 [47 to 66] years. Patients were randomized to anakinra (42, 67%) or placebo (21, 33%). Most of the clinical characteristics were not significantly different between the two groups, except sex with more males represented in the placebo group, higher body mass index (BMI) values in the anakinra group, and the VE/VCO_2_ slope, higher in placebo (**Table 1**). CRP levels were 6.7 [3.0 to 14.0] mg/L, and VO_2peak_ was 13.0 [11.8 to 17.2] mL·kg^-1^·min^-1^, corresponding to an absolute VO_2peak_ of 1.29 [1.08 to 1.53] L/min, with no significant differences between groups. The longest available follow up was 6 weeks in 10 patients (16%), 12 weeks in 12 patients (19%), and 24 weeks in 41 patients (65%) with a median of 24 [12 to 24] weeks.

### Effect of treatment on the IL-1Ra Levels

Biomarker and functional data at baseline and follow-up are reported in **Table 2**. IL-1Ra levels were significantly higher in patients treated with anakinra compared with placebo, with values of 3,994 [3,372 to 5,000] pg/mL versus 492 [304 to 1,370] pg/mL, respectively (*P*<0.001, **Table 2**). The interval change in IL-1Ra from baseline to the on-treatment assessment was also greater in the anakinra group, increasing by 3,374 [1932 to 3961] pg/mL compared with 33 [−60 to 600] pg/mL in the placebo group (*P*<0.001, **Table 2**). Similar results were observed across the different follow-up time points, with significantly higher IL-1Ra levels and greater interval changes in the anakinra group at each available follow-up assessment (*P*<0.05, **Figure 2**).

**Figure 2.**
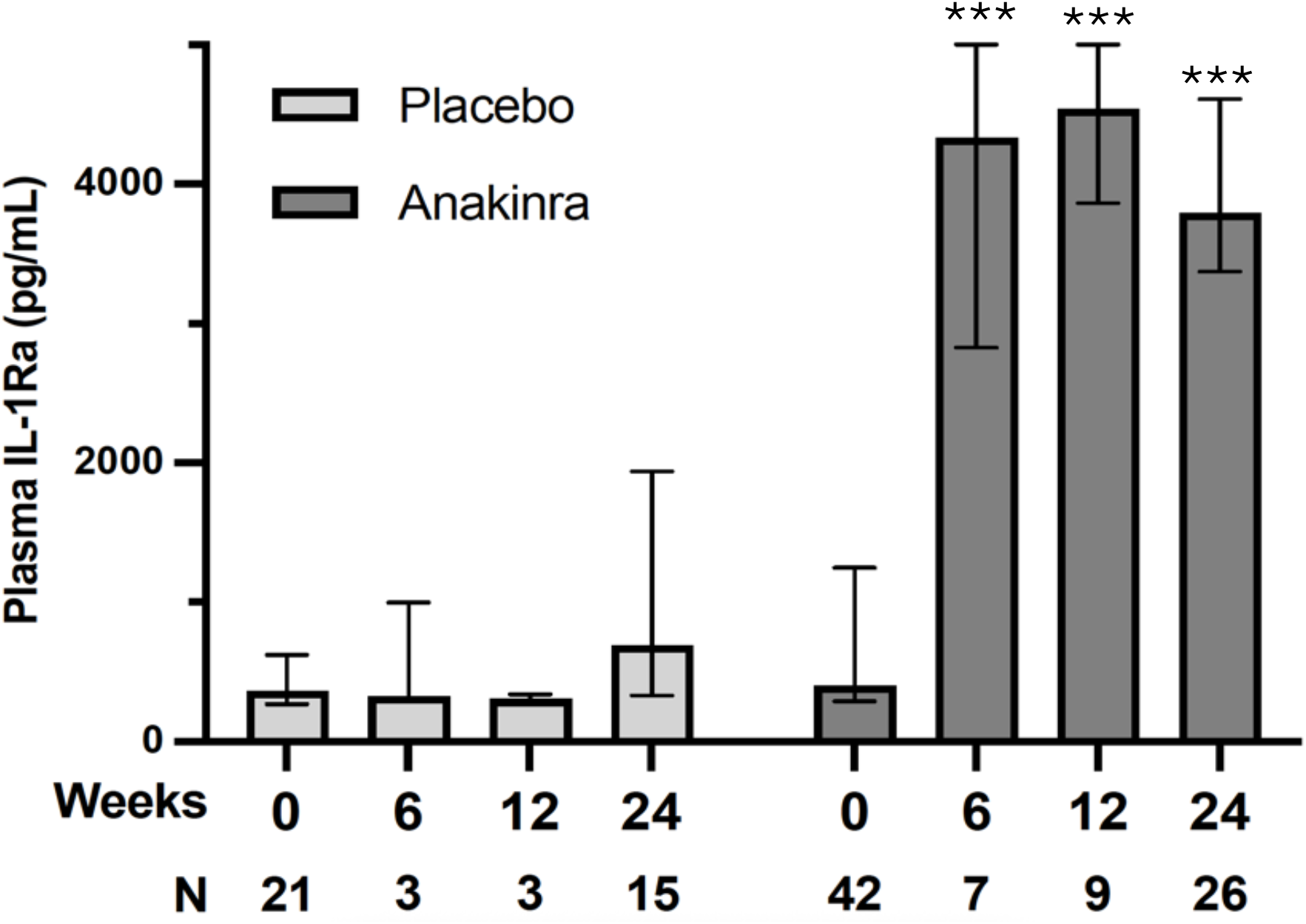
Plasma IL-1Ra levels across follow-up time points. Plasma Interleukin-1 receptor antagonist (IL-1Ra) concentrations (pg/mL) are shown at baseline (week 0) and at follow-up time points (weeks 6, 12, and 24) in patients randomized to placebo (light gray) or anakinra (dark gray). Data are presented as median with interquartile range. A marked increase in IL-1Ra levels was observed in the anakinra group compared with placebo, consistent across all follow-up time points. Sample sizes at each time point are indicated below the x-axis. \*\*\**P*<0.001 vs. placebo at the corresponding time point.

No statistically significant differences in IL-1Ra levels were observed between male and female participants, and no significant correlation was found between IL-1Ra levels and BMI or VE/VCO_2_ slope (data not shown).

### Effect of IL-1Ra levels on CRP and Peak VO2

At the last available follow-up assessment, CRP levels were 3.3 [0.8 to 10.1] mg/L in the placebo group and 1.71 [0.6 to 3.9] mg/L in the anakinra group (*P*=0.15, **Table 2**). VO_2peak_ values were 15.3 [12.4 to 17.9] mL·kg^-1^·min^-1^ and 1.50 [1.13 to 1.72] L/min in the placebo group, and 15.0 [12.1 to 18.5] mL·kg^-1^·min^-1^ and 1.52 [1.22 to 2.02] L/min in the anakinra group (*P*=0.39 and *P*=0.39, respectively, **Table 2**). IL-1Ra levels and interval change in IL-1Ra showed a modest inverse correlation with CRP levels (R=−0.269, *P*=0.033 and R=−0.355, *P*=0.004, respectively; **Figure 3**). In contrast, no statistically significant correlations were observed between on-treatment IL-1Ra levels or change in IL-1Ra and VO_2peak_ values, whether assessed as absolute values or as interval changes over time (all *P*>0.05).

**Figure 3.**
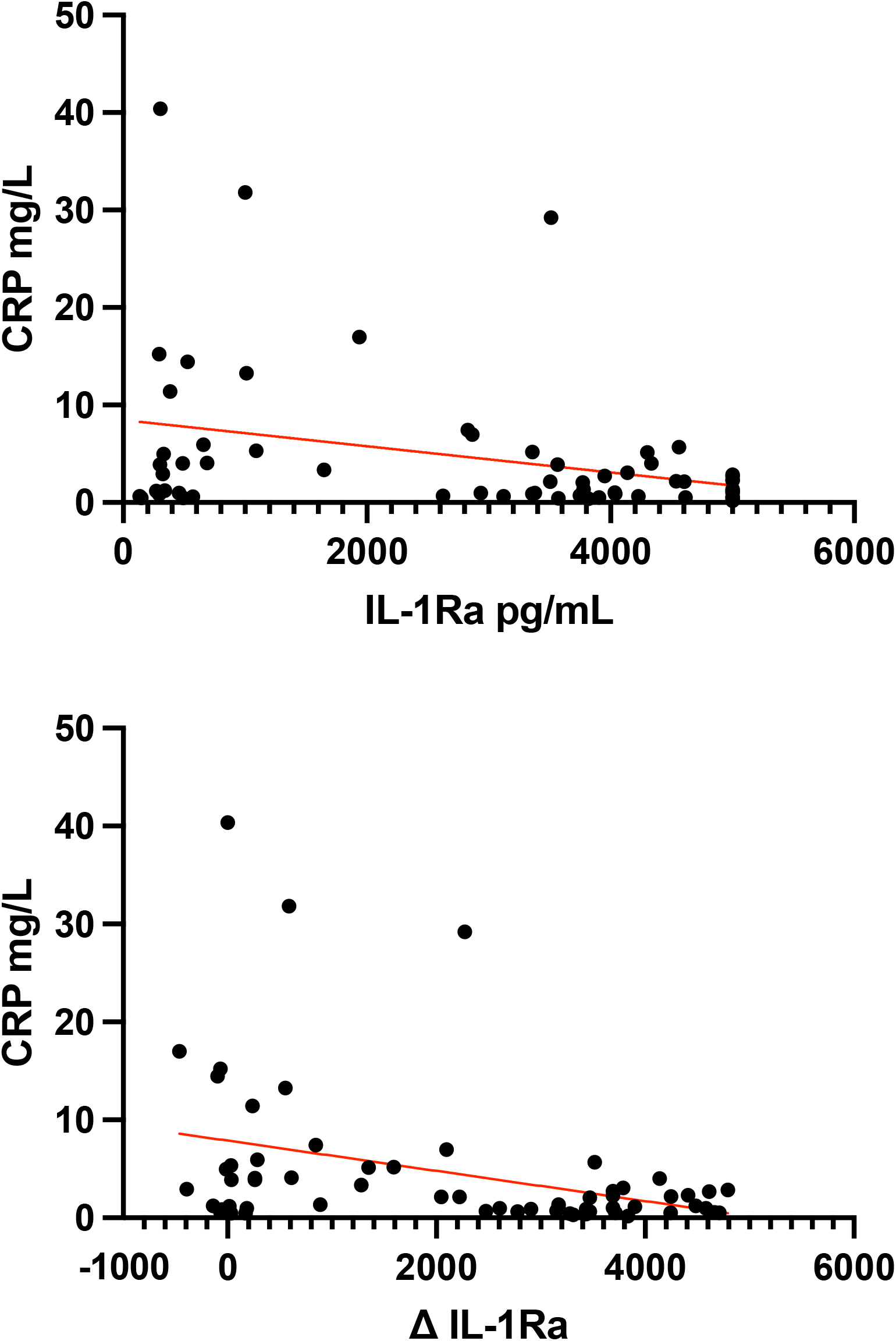
Correlation between on-treatment IL-Ra levels and CRP levels. Plasma Interleukin-1 receptor antagonist (IL-1Ra) concentrations (pg/mL) were measured at baseline and follow up. The top panel shows correlation between on-treatment IL-1Ra levels and C-reactive protein (CRP) levels (R=−0.269, *P*=0.033). The bottom panel shows correlations between changes in IL-1Ra levels (ΔIL-1Ra) and on-treatment CRP levels (R=−0.355, *P*= 0.004).

## DISCUSSION

In this substudy of patients with recently decompensated heart failure with reduced ejection fraction (HFrEF) enrolled in the REDHART2 trial, treatment with anakinra resulted in a significant increase in circulating IL-1Ra levels. Anakinra is a recombinant form of the naturally occurring IL-1Ra that acts by competitively inhibiting IL-1 signaling.^13^ Patients receiving anakinra exhibited substantially higher on-treatment IL-1Ra concentrations than those assigned to placebo, due to a greater interval increase from baseline. These findings confirm that administration of anakinra achieved the expected pharmacological effect of the drug.

Importantly, this increase was consistently observed across the different available follow-up time points, indicating a robust and sustained biological response to treatment and suggesting that adequate systemic drug exposure was achieved in most treated patients (**Figure 2**).

In addition to confirming anakinra’s pharmacologic activity, our analysis provides insight into its biologic activity exploring the relationship between IL-1 pathway inhibition and systemic inflammation. We observed a modest but significant inverse correlation between on-treatment IL-1Ra levels and CRP concentrations, as well as between the interval changes in IL-1Ra and CRP levels at follow-up (**Figure 3**). These findings suggest that increased exposure to IL-1Ra is associated with a greater reduction in systemic inflammation. This correlation between CRP and IL-1Ra was, however, only moderate in strength and many patients failed to normalize their CRP levels despite the increased IL-1Ra levels. This suggests that either the levels of IL-1Ra levels were not high enough or that there are other IL-1-independent signals leading to systemic inflammation.

Moreover, despite anakinra’s biological effect on IL-1Ra levels and its moderate association with CRP reduction, we did not observe a significant relationship between IL-1Ra levels and measures of cardiorespiratory fitness, including VO_2peak_. In the present analysis, VO_2peak_ was assessed both as absolute oxygen consumption (L/min) and normalized to body weight. No significant association was observed between on-treatment IL-1Ra levels and VO_2peak_, whether VO_2peak_ was analyzed using absolute or relative values at follow-up or as the interval change from baseline. These findings suggest that, although IL-1 blockade effectively modulates inflammatory activity, the resulting changes in systemic inflammation did not translate into measurable improvements in functional capacity within the timeframe of the present study.

These results are consistent with the main REDHART2 trial results, which showed only a modest additional effect of anakinra on inflammatory markers and no significant improvement in VO_2peak_ or other measures of cardiorespiratory fitness compared with placebo.^11^ As discussed in the initial study, patients received contemporary guideline-directed medical therapy and were closely monitored during follow-up, leading to improvements in both CRP levels and VO_2peak_ even in the placebo group. Such optimization of heart failure therapy and intensive follow-up may have reduced residual inflammatory risk and limited the ability to detect additional functional benefits of IL-1 blockade. ^11^

Nevertheless, inflammation may still be an important therapeutic target in cardiovascular disease. In the CANTOS trial, inhibition of IL-1 with canakinumab reduced recurrent cardiovascular events and heart failure hospitalizations in patients with prior myocardial infarction and elevated inflammatory risk.^4^ Similarly, recent meta-analyses have shown that anti-inflammatory therapies can reduce systemic inflammatory markers and may improve clinical outcomes in selected populations.^14^ Therefore, it is plausible that the magnitude or duration of IL-1 inhibition achieved with the dosing strategy used in this trial was insufficient to fully suppress inflammatory signaling in this population. Future studies may need to explore higher doses of IL-1 blockade, alternative dosing regimens, or the use of more potent and/or longer-acting anti-inflammatory agents.

This study has several limitations, including a relatively small sample size that limits the statistical power of the analysis and the ability to detect modest associations between biological exposure to IL-1 blockade and functional outcomes. Moreover, follow-up measurements of IL-1Ra levels were not available for all patients at the final study visit, and not all participants underwent biomarker assessment at visit 4 (24 weeks), resulting in variable follow-up duration across the study population.

## CONCLUSION

In conclusion, treatment with anakinra in patients with recently decompensated HFrEF resulted in a marked increase in circulating IL-1Ra levels, confirming an effective pharmacological profile. Higher IL-1Ra levels were associated with lower CRP concentrations, supporting the known biological link between IL-1 blockade and reduction of systemic inflammation. However, the association with CRP was only modest, and no association was observed between IL-1Ra levels and cardiorespiratory fitness. These findings highlight the complexity of the relationship between inflammatory signaling and functional capacity in HFrEF and support further investigation of anti-inflammatory strategies in carefully selected patient populations with persistent inflammatory activation.

## Data Availability

All data produced in the present study are available upon reasonable request to the authors

